# Impact of a multimodal hand hygiene improvement intervention in a 1000-bed hospital in Thailand: a stepped wedge cluster randomized controlled trial

**DOI:** 10.1101/2022.11.24.22282731

**Authors:** Maliwan Hongsuwan, Pramot Srisamang, Somboon Nuntalohit, Nantasit Luangasanatip, Cherry Lim, Nicholas P. Day, Direk Limmathurotsakul, Ben S. Cooper

## Abstract

**Background:** Good hand hygiene compliance amongst healthcare workers is critical for patient safety and plays a central role in preventing healthcare-associated infections. The World Health Organization (WHO) recommends a multimodal strategy to improve healthcare worker hand hygiene. We aimed to evaluate the effectiveness of this strategy in a middle-income country using a stepped-wedge cluster randomized trial.

**Methods:** The trial was conducted between 2013 and 2015 in 58 wards in a 1000-bed hospital in Thailand. The intervention was adapted from the WHO’s Hand Hygiene Improvement Strategy and implemented by the hospital’s infection control team. The primary outcome was observed hand hygiene compliance among healthcare workers in the study wards. This study was registered at ClinicalTrials.gov (NCT01933087).

**Findings:** During 4,230 observation sessions, 54,073 hand hygiene opportunities were identified. Hand hygiene compliance increased from 10.0% (2,660/26,482) to 11.0% (3,048/27,591) after the intervention (odds ratio [OR] 1.12; 95% CI: 1.01-1.24). Among the five moments for hand hygiene, the greatest improvement in compliance was observed in hand hygiene before patient contact (OR 1.52; 95% CI: 1.21, 1.91).

**Interpretation:** While hand hygiene compliance improved slightly, the intervention failed to achieve the substantial improvements that were needed. There is a need for new strategies to ensure that all hospitals in low and middle-income countries can achieve and maintain acceptable levels of hand hygiene.

**Funding:** Oak Foundation, MRC

## Introduction

Healthcare-associated infections (HCAIs) are a major source of preventable morbidity and mortality worldwide, and particularly so in low- and middle-income countries (LMICs).^1-3^ In 2011 the World Health Organization (WHO) estimated that 7% of patients in developed and 10% of patients in developing countries acquired at least one HCAI during hospital admission.^2,4^ Mortality due to such infections in developing countries exceeds that in developed countries.^5^

Improving hand hygiene practice among healthcare workers (HCWs) can substantially reduce the transmission of important healthcare-associated pathogens resulting in reduced incidence of HCAI and is a key intervention for combating multidrug-resistant (MDR) bacterial infections in hospitals.^6,7^ Successful hand hygiene campaigns are associated with reductions in incidence rates of MDR bacterial infections at the hospital, regional and national levels. Interventions to improve hand hygiene behavior are relatively inexpensive and expected to be cost-effective (and potentially cost-saving) in most circumstances.

The WHO established evidence-based guidelines on hand hygiene in healthcare settings to support efforts to improve hand hygiene compliance.^8^ These guidelines, which were designed to be used in all healthcare facilities, irrespective of the level of resources or prior hand hygiene initiatives, advocate a multimodal strategy consisting of five components: 1) system change, 2) training and education, 3) observation and feedback, 4) reminders in the work place, and 5) a hospital safety climate.^9^ A recent systematic review concluded that the WHO multimodal hand hygiene improvement strategy is effective at improving HCW hand hygiene compliance.^10,11^ However, evaluations of the effectiveness of this strategy in LMICs using methodologically robust study designs were found to be lacking.^4,10,12^ A more recent systematic review of interventions to improve hand hygiene compliance in patient care concluded that there is “an urgent need to undertake methodologically robust research to explore the effectiveness of multimodal versus simpler interventions to increase hand hygiene compliance, and to identify which components of multimodal interventions or combinations of strategies are most effective in a particular context”.^13^ Given that LMICs face the highest burden of HCAIs and also unique challenges related to delivery of effective infection control interventions, we sought to address this research gap.

A stepped wedge design (where all clusters receive the intervention but at randomly allocated times) is appropriate when there are prior reasons to believe the intervention will be beneficial, as was the case here.^14-16^ Although hand hygiene promotion is part of routine work for infection control teams (ICTs) in most provincial hospitals in Thailand, hand hygiene levels have been consistently reported to be below 45% in hospitals. ^17-19^ The aim of this study was to evaluate the impact of a multimodal intervention to improve hand hygiene compliance amongst HCWs in a LMIC using a stepped wedge cluster-randomized trial.

## Methods

### Study design, setting and Participants

A stepped wedge cluster randomized controlled trial was performed in a resource-limited tertiary hospital in northeast Thailand between 1^st^ December 2013 and 2^nd^ May 2015. Fifty-eight wards, which included 20 intensive care units (ICUs) and 38 non-ICU wards were included in this study. The trial included a 5-week baseline period (phase 1), a 64-week intervention period (phase 2), and a 5-week follow-up period (phase 3) (Figure 1).

**Figure 1:**
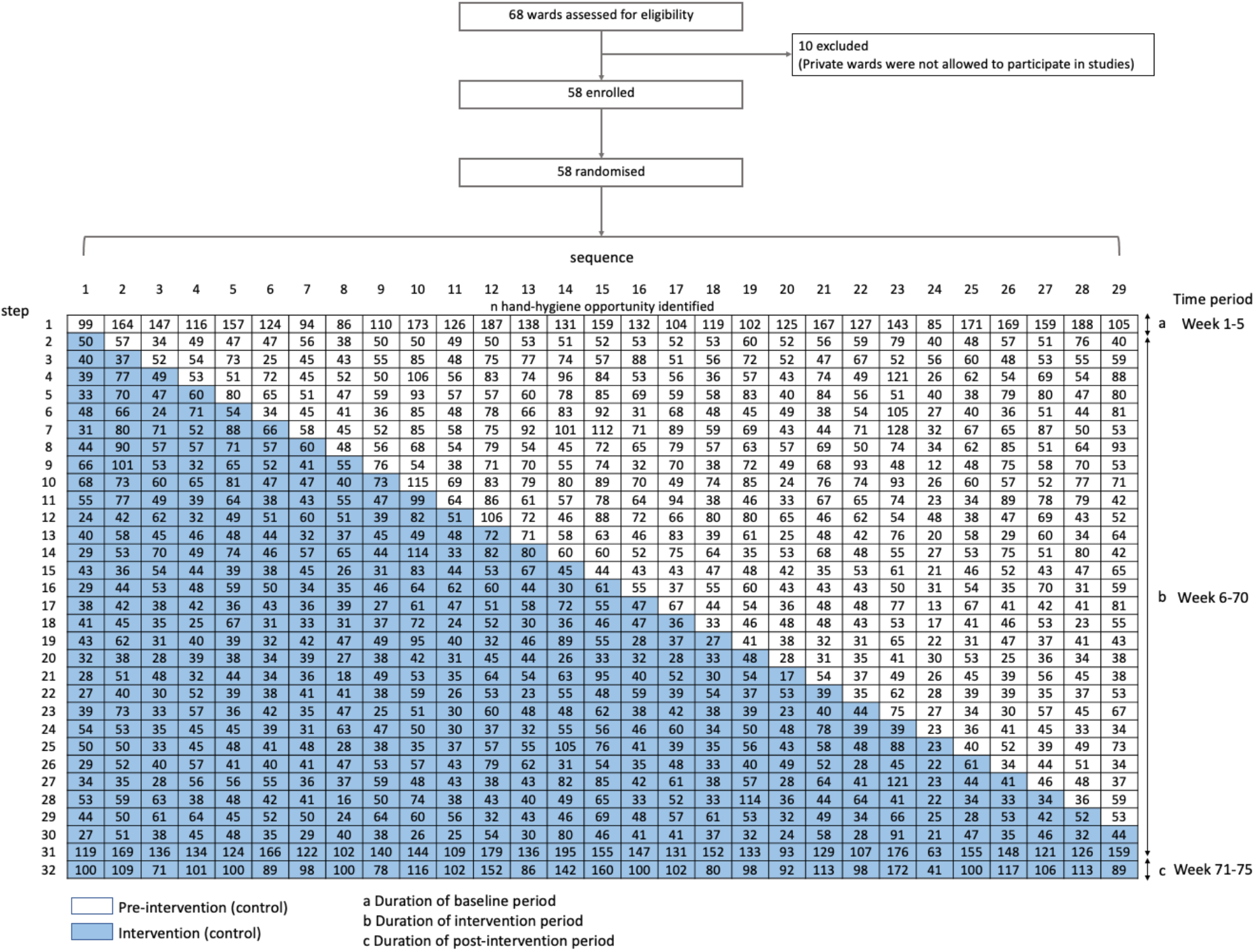
Design of the stepped-wedge cluster randomized trial.

During the study period, the hospital had 1000-beds, and an ICT consisting of four infectious disease physicians, four specialist infection control nurses, and one assistant. Hospital-produced alcohol-based hand sanitizer was readily available in all wards throughout the study period.

All hospital staff who had direct patient contact were eligible to be included in the study. The participants included physicians, nurses, nursing aides and hospital support staff who worked in the participant wards on a regular basis. Other hospital staff who were included in the study and had direct contact with patients but did not work in the study wards on a regular basis were medical students, nursing students, physical therapists, and radiologists. In 2014, the four members of the ICT attended a training course on “Healthcare-associated Infection Prevention and Management” held by ESCMID-SHEA Training Course in Hospital Epidemiology. The ICT had performed infection control training for all HCWs annually, and monitored hospital preventive measures during hospital accreditation events and when an outbreak was detected.

This study was approved by the Ethical and Scientific Review Committees of Faculty of Tropical Medicine, Mahidol University, Thailand (MUTM 2012-042-01), and Sunpasitthiprasong Hospital (002/2556). This study was registered at ClinicalTrials.gov (NCT01933087).

### Randomization and procedure

After a 5-week baseline period, fifty-eight participating wards were randomized to one of 29 intervention dates which occurred at two-week intervals (Figure 2). Randomization was performed by a statistician using a computer-generated sequence with two wards randomly selected to receive the intervention on each of the intervention dates. Masking of study participants and observers was not possible due to the nature of the interventions.

**Figure 2:**
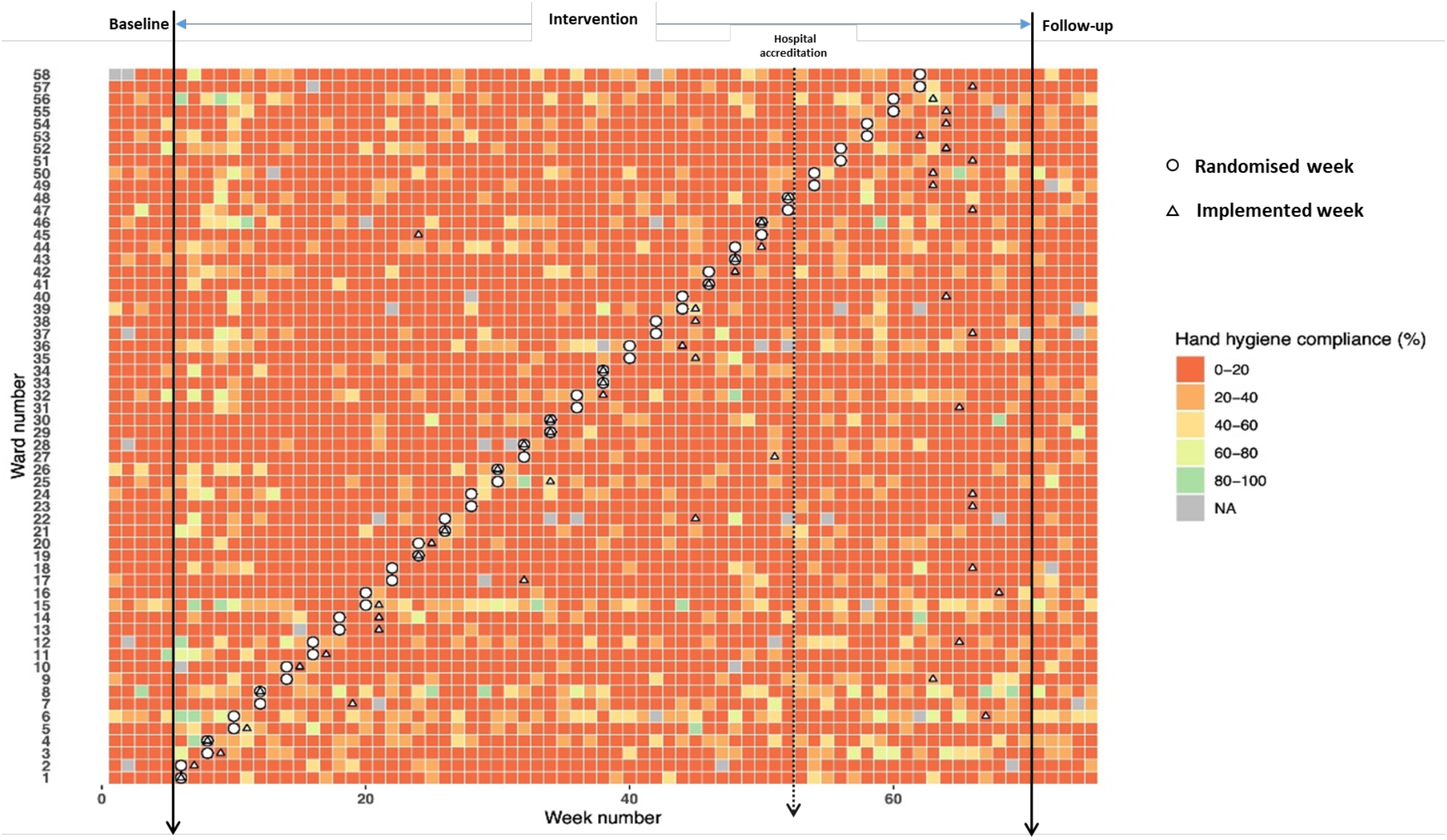
Hand-hygiene compliance over the study period.

### Study intervention

The study intervention was derived from the WHO Multimodal Hand Hygiene Improvement Strategy. ^20^ In order to engage ward staff in the decision-making process, to increase their ownership of the intervention, and to ensure the intervention was adapted to local conditions in each ward, the implementation details of the intervention were chosen by ward staff in consultation with the ICT. To do this a set of actions was presented to ward staff as a “five-course menu” with each “course” representing one of the five elements of the WHO multimodal strategy. Staff on each ward were asked to create a definite plan to implement all the strategy components by choosing at least one item from each “course”. Ward staff were also encouraged to create original ideas for improving the implementation of the strategy components. The intervention package, which was provided in the local (Thai) language in both the electronic and hard copy format, contained four key elements: (1) details of each component of the WHO strategy with a number of suggestions for how this could be applied and a minimum requirement for implementing each component; (2) details of the step-wise approach, which determined the intervention steps; (3) details of WHO recommendations for implementation of each component of the WHO strategy; and (4) ward self-assessment that allowed the ward to consider all components of the strategy.

In addition, staff on each ward were asked to follow the step-wise approach of the WHO Multimodal Hand Hygiene Improvement Strategy, which included 1) facility preparedness, 2) baseline evaluation, 3) implementation, 4) follow-up evaluation, and 5) ongoing planning and review cycle as part of the implementation process.^20^

### Outcome measurements

The primary outcome was compliance with the WHO’s recommended five moments for hand hygiene amongst HCWs who had direct patient contact. This was assessed through direct observation by the investigation team. Inter-rater reliability was assessed before and during the study on a monthly basis with hand hygiene compliance data recorded independently by different observers for the same hand hygiene opportunities.

The trial start and finish date were pre-specified as 24^th^ December 2013 and 2^nd^ May 2015. Directly observed hand hygiene compliance data were collected during the study by observers. Staff were not informed that their hand hygiene behavior was being observed. The protocol specified 58 observation sessions (one per ward), each lasting 15-30 minutes, in each week of the study, with a minimum of five hand hygiene opportunities observed in each session.

The observers were trained to record hand hygiene compliance data in accordance with WHO recommendations, using tools for training and education for observers provided by the WHO (http://www.who.int/gpsc/5may/tools/training_education/en/) and also making use of the WHO’s “5-movements” framework and the standard WHO hand hygiene compliance observation forms.^9^ Hand hygiene observation data were entered daily on a secure password-protected study database implemented in OpenClinica®.

### Sample size calculation

Power calculations were performed based on Hussey & Hughes (2007).^21,22^ Assuming 52 wards, and 60 one-week time periods with the observation of an average of five hand hygiene opportunities per ward per week throughout the study, and with an intra-cluster correlation coefficient of 0.01 and baseline compliance 5%, the power to detect a 2% increase in hand hygiene significant at the 0.05 level was 93%; this reduced to 70% for a baseline compliance of 10%. We assumed extra binomial variation in hygiene compliance increased the outcome variance by a factor of two.

### Statistical analysis

The primary analysis was done on an intention to intervene basis (using the weeks at which each ward was randomized to receive the intervention). Response functions for modelling the effect of the intervention allowed for both changes in level and trend of outcome measures. The analysis was performed at the cluster level, with observations of hand hygiene compliance within each ward for each time period (week of study) and used a generalized linear mixed model. Analysis was performed using STATA version 14.0 (StataCorp LP, College station, Texas). The overall compliance for each ward in each week was calculated as the average compliance over all observed hand hygiene opportunities. An additional per protocol analysis (using the weeks when wards actually received the intervention) was performed using the same multilevel logistic regression analysis accounting for clustering at the ward level. Exploratory subgroup analyses were performed for hand hygiene opportunities corresponding to each one of the “five moments” and by ward type. Cohen’s kappa statistic was used to quantify inter-rater reliability between the observers.^23^ A sensitivity analysis to relax the assumption of immediate treatment effect was performed as described elsewhere.^22^ In brief, exposure time (the amount of time that has passed since the start of the treatment as assigned for a given cluster) was included in the model.

## Results

Intention-to-treat analysis was performed on the 58 study wards using the assigned intervention implementation date. Thirty-two wards (57%) initiated implementation of the intervention within one month of the time specified at randomization. Twenty-three wards (40%) had a delay greater than one month (range 5−57 weeks). One ward implemented the intervention before the week of randomization due to an administrative error. One ward refused to participate in the study and one ward failed to implement the WHO multimodal strategy during the intervention period. Intention-to-treat analyses were performed on 57 participating wards, and per-protocol analyses were performed on the 56 wards using the actual week of implementing the intervention (Figure 1). To ensure that the last ward that implemented the intervention had a 5-week period of post-intervention data collection, the data collection period was extended for all wards by six weeks.

### Hand hygiene compliance observations

Inter-rater reliability of hand hygiene observations (based on 18 assessments made throughout the study) was good and the mean kappa score was 0.93. There were 4,230 observation sessions. In total, 54,073 hand hygiene opportunities were observed throughout the study (Table 1). The median number of observed opportunities per ward per week was 11 (IQR 8-16).

**Table 1:** Hand hygiene opportunities during baseline, intervention, and post-intervention periods.

|  | Baseline | Intervention | Follow-up |
| --- | --- | --- | --- |
| n | 3,907 | 47,141 | 3,025 |
| Day |  |  |  |
| Monday | 772 (19.8%) | 8,628 (18.3%) | 600 (19.8%) |
| Tuesday | 835 (21.4%) | 8,265 (12.5%) | 483 (16.0%) |
| Wednesday | 839 (21.5%) | 7,577 (16.1%) | 577 (19.1%) |
| Thursday | 668 (17.1%) | 11,996 (25.4%) | 612 (20.2%) |
| Friday | 682 (17.5%) | 9,466 (20.1%) | 647 (21.4%) |
| Saturday | 111 (2.8%) | 512 (1.1%) | 106 (3.5%) |
| Sunday | 0 (0%) | 697 (1.5%) | 0 (0%) |
| Time of day |  |  |  |
| Morning | 2248 (57.5%) | 24,517 (52.0%) | 1,673 (55.3%) |
| Afternoon | 1659 (42.5%) | 22,624 (48.0%) | 1,352 (44.7%) |
| WHO Moment |  |  |  |
| 1-before touching a patient | 859 (22.0%) | 10,580 (22.4%) | 621 (20.5%) |
| 2-before clean or aseptic procedure | 1068 (27.3%) | 10,210 (21.7%) | 375 (12.4%) |
| 3-after body fluid exposure risk | 444 (11.4%) | 5,911 (12.5%) | 320 (10.6%) |
| 4-after touching a patient | 691 (17.7%) | 6,594 (14.0%) | 2279 (9.2%) |
| 5-after touching patient surroundings | 845 (21.6%) | 12,846 (29.4%) | 1,430 (47.3%) |
| Profession |  |  |  |
| Physicians | 291 (7.4%) | 2,223 (4.7%) | 60 (1.7%) |
| Nurses | 2,280 (58.4%) | 28,591 (60.6%) | 2,169 (71.7%) |
| Auxiliaries | 884 (22.6%) | 12,261 (26.0%) | 737 (24.4%) |
| Nursing students | 36 (0.9%) | 403 (0.9%) | 0 (0%) |
| Medical students | 188 (4.8%) | 2,212 (4.7%) | 69 (2.3%) |
| Visiting specialist | 228 (5.8%) | 1,451 (3.1%) | 0 (0%) |

Data are n (%), unless indicated otherwise.

During the baseline period (phase 1), 3,907 hand hygiene opportunities were observed with a median compliance per observation session of 0% (IQR 0-10%). During the intervention period (phase 2), 47,141 hand hygiene opportunities were observed with a median compliance of 5% (IQR 0-17%), while for the follow-up period (phase 3), 3,025 hand hygiene opportunities were observed with a median compliance of 0% (IQR 0-17%).

More than half of the observed hand hygiene opportunities came from nurses (61.1% [33,040/54,073]), followed by auxiliaries (25.7% [13,882/54,073]), and physicians (4.7% [2,564/54,073]). Thirty-eight of 57 wards were non-ICUs. These wards accounted for the majority of observed hand hygiene opportunities (68.5% [37,050/54,073]).

### Overall impact of the intervention on hand compliance

Hand hygiene compliance in the pre- and post-intervention periods, as defined per actual implementation, was 10.4% (3,384/32,634) and 11.0% (3,048/27,591), respectively (Figure 2). The intention-to-treat analysis indicated a small effect of the intervention on hygiene compliance (odds ratio [OR] 1.10; 95% confidence interval [CI]: 1.00-1.21) (Table 2). After adjustment for time of day, day of the week, month of the year, type of ward (ICU or non-ICU), and type of HCW, the estimated OR was 1.11 (95% CI: 1.00-1.22). An interaction term was added to account for the different magnitudes of effect of intervention on compliance by type of ward. The largest impact of the intervention was in the department of obstetrics and gynecology (OR 3.96; 95% CI 1.88, 8.31). Per-protocol analysis showed similar findings and indicated a small improvement in hand hygiene compliance (Table 2). A sensitivity analysis that relaxed the assumption of immediate treatment effect by including the exposure time in the model showed similar findings.

**Table 2:**
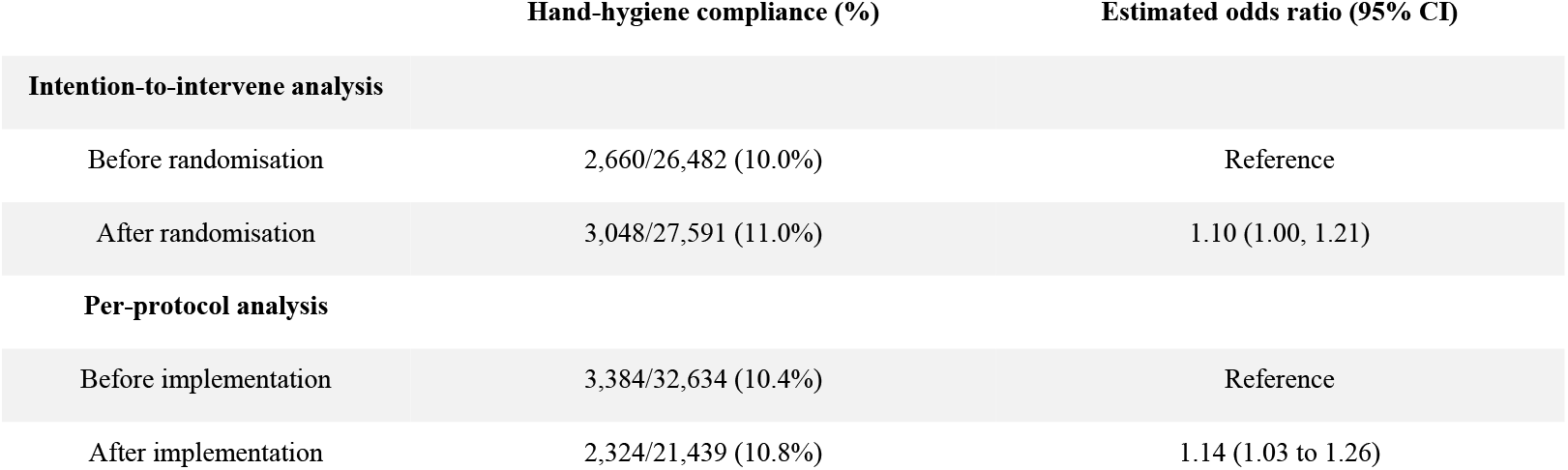
Estimated effect of intervention on hand hygiene compliance.

### Hand hygiene compliance stratified by “My five moments for hand hygiene”

The most commonly observed moment for hand hygiene was ‘after touching patient surroundings’, which accounted for 30.2% (16,121/54,073) of all hand hygiene opportunities, followed by ‘before touching a patient’, which accounted for 22.6% (12,060/54,073) (Table 1). The highest effects of the intervention were found for the indication ‘before touching a patient’ and the estimated odds ratio from the intention-to-treat analysis was 1.52 (95%: CI 1.21, 1.91). Per-protocol analysis gave similar findings.

### Variation in hand hygiene compliance by ICU ward and type of ward staff

Hand hygiene compliance in ICU wards (15.4% [2,624/17,023]) was higher than in non-ICU wards (8.3% [3,084/37,050]). Compliance in non-ICU wards increased from 7.0% before intervention to 9.4% of after intervention, while in ICU no improvements in compliance were seen (15.6% before intervention vs. 15.1% of after intervention in the ITT analysis).

Hand hygiene compliance varied by the type of HCW during the pre-intervention period. The highest compliance was observed in nurses (10.0% [229/ 2,280]), followed by that in medical students (6.9% [13/188]), and then auxiliaries (5.4% [48/884]). The highest absolute improvement in hand hygiene compliance was seen in student nurses (3.3% before intervention vs. 6.2% after interventions), followed by physicians (7.3% before intervention vs. 9.6% of after intervention), and nurses (11.4% before intervention vs. 12.6% of after intervention).

## Discussion

The principal finding is that while a multimodal intervention based on the WHO’s Hand Hygiene Improvement Strategy led to improvements in hand hygiene compliance, the improvements were small and would not be expected to substantially reduce HCAI rates. To our knowledge, this is the first time a hospital-wide implementation of the WHO hand hygiene strategy has been evaluated in a randomized trial in a LMIC, so it is important to consider why it was unsuccessful at achieving the desired change. We believe three main factors are likely to have been important. First, human resource limitations are likely to have played a role. Because we wanted our findings to be generalizable to other resource-limited hospitals in LMICs, the intervention only made use of resources normally available to the hospital and its infection control team. We therefore deliberately did not employ additional infection control staff for delivering the intervention; nor did we bring in external expertise in behavior change or social marketing, and all promotional and training materials were produced in-house without professional help. We did, however, provide access to high quality training in infection control to members of the hospital infection control team before the intervention started, with members of the ICT attending a course on “Healthcare-associated Infection Prevention and Management” held by ESCMID-SHEA Training Course in Hospital Epidemiology in 2014. We also translated the WHO’s hand hygiene strategy into Thai (https://doi.org/10.21954/ou.ro.0000d7f3), to ensure that infection control staff were able to acquire a good knowledge of the WHO strategy. We provided additional resources for evaluating the intervention to ensure that this did not in any way impact on the ICT’s workload. Nonetheless, the ICT were not always able to deliver the intervention as planned because of competing infection control and organizational priorities including routine surveillance, responding to outbreaks and preparing for a nationally mandated hospital accreditation event. While we did not attempt to assess qualitative aspects of the intervention components which were implemented, clearly some approaches to providing reminders in the workplace, training, system change, promoting a safety climate, and giving feedback are likely to be more effective than others. We can speculate that had more resources, including time and professional expertise in designing effective behavior change interventions, been devoted to these components there would have been more chance of effecting meaningful improvements.

Second, while certain wards did take ownership of the intervention as had been the intention, resulting in some substantial ward-specific improvements in hand hygiene behavior, a more common pattern was that the intervention was not fully implemented or was implemented late. The ICT were not always able to obtain the co-operation of the majority of ward staff despite high-level endorsement of the intervention by the hospital director. This reflects the fact that although the intervention was explicitly endorsed by senior hospital management, organizational factors and other priorities sometimes got in the way. For example, hospital staff changes occurring during the study caused some delays in implementing the intervention. Other studies have reported similar problems, with higher priority work preventing effective implementation of hand hygiene interventions.^24^

Third, physical resource limitations may have also played a role, particularly for opportunities when hand washing with soap and water was indicated, and insufficient provision of conveniently located sinks and lack of provision of towels at sinks were both reported by hospital staff as likely contributory factors to poor hand hygiene compliance (https://doi.org/10.21954/ou.ro.0000d7f3)

The baseline hand hygiene compliance in this study of about 10% is low, but not atypical. Similar pre-intervention rates have been reported in other studies in both low- and high-income countries.^25-27^A recent systematic review found compliance rates of below 20% to be common for all types of HCW.^28^

The most extensive previous evaluation of the WHO’s multimodal strategy for improving hand hygiene in LMIC settings used a before-after study design and included three countries: Mali, Costa Rica and Pakistan.^27,29^ Substantial improvements in hand hygiene in all three countries were reported: respectively, from 8% pre- to 22% post-intervention compliance in Mali, from 40% to 60% in Costa Rica, and from 38% to 59% in Pakistan. Although the before-after design employed in this study is considered to be weak because of its vulnerability to secular trends, the magnitude of the changes reported is consistent with moderate to large beneficial effects. In a similar manner to our study, the interventions were obtained mainly with the use of local resources and expertise. One important difference, however, is the direct involvement of the WHO team who developed the multimodal strategy. Assuming that the changes seen were caused by the intervention, it is possible that the involvement of such a high-profile team helped to galvanize behavior change amongst hospital staff and overcome organizational inertia. More recently, one other study in Sub-Saharan Africa has reported substantial increases in hand hygiene compliance (from 34% at baseline to 69% post-intervention), but again using a weak before-after study design. Assessment of the published literature also needs to take into account the likelihood of publication bias, whereby small (often methodologically weak) study designs are more likely to be reported if they show a positive result.^13,30^ A previous systematic review of hand hygiene interventions that included such before-after studies found clear evidence of such publication bias,^30^ and it is notable that large studies of hand hygiene interventions employing the strongest study designs (and with pre-registered protocols) have consistently shown the smallest effects.^31-33^ Indeed, our estimated effect size is consistent with those seen in previous interrupted time series studies.^31^ For these reasons, we believe that our experience of an intervention that achieved a statistically but not clinically significant improvement in hand hygiene may be representative of an underlying problem that has been largely neglected from the published literature. Indeed, the difficulties in implementing the intervention as planned in this study echo those from a previous CRCT of a hand hygiene improvement intervention,^34^ which found that poor fidelity to the planned intervention may have substantially diluted the potential benefits. Competing priorities with other quality improvement initiatives were identified as one possible cause of these implementation difficulties.

Well-resourced national campaigns modelled on the WHO strategy have helped to bring large, persistent and well-documented improvements in hand hygiene compliance in some high-income countries and these have been shown to be associated with reductions in infection.^35,36^There is also some evidence that the WHO strategy can be further improved by adding interventions including goal-setting, reward incentives, and accountability.^10^ In our study, goal-setting was used as an additional intervention by two wards who chose to have routine meetings to evaluate the intervention plan, revise the goals and plan to achieve target rates of hand hygiene. It has also been proposed that team–based approaches could help promote hand hygiene with ward-based workshops to discuss the workflow and make ward-specific plans.^24^ Using this approach Huis *et al*. found high improvement in certain wards that were successful in making culture changes at the ward to make staff more aware of performing hand hygiene.^37^ In our study, there were also examples of creative ideas originating from ward staff to promote hand hygiene. For example, some wards set a hand hygiene day in every month. On these days there were activities to promote hand hygiene in the ward in a similar manner to the annual hospital hand hygiene day.

The principal strength of our study is the strong design used to evaluate the impact of the WHO strategy,^16,34^ and the hospital-wide intervention. Most previous studies from LMICs have evaluated interventions only in more restricted study populations.

The study’s main limitation was that, as with other hand hygiene studies based on direct observation, Hawthorne effects may have distorted HCW hand hygiene behavior. We had hoped to overcome this problem by independently assessing hand hygiene compliance by measuring volume of alcohol-based hand sanitizer use on each ward, though this proved impossible to do reliably in practice. A second limitation is the possibility of contamination between clusters, as interventions made on one ward could affect staff behavior on other wards. This would be expected to result in overall increases in compliance over time, but might lead to smaller differences between pre- and post-intervention periods in implementing wards.

## Conclusions

Hand hygiene is an essential part of infection control practice and there is compelling evidence that it can be highly effective at reducing nosocomial infection, particularly MDR bacterial clones adapted for hospital transmission. Previous studies in high income settings have had remarkable success in substantially improving hand hygiene in HCWs using interventions based on the WHO’s multimodal strategy. Such successes have been documented at the level of the hospital, at a regional level, and nationally. Each time the improvements have been accompanied by substantial reductions in multi-drug resistant infections. However, with the exception of countries, such as Australia, which require independent auditing and public reporting of hand hygiene compliance in all hospitals, we do not have a good idea of how widespread these improvements have been nor have there been methodologically rigorous evaluations of the WHO’s multimodal strategy in resource-constrained settings. Our study, performed in a typical resource-constrained hospital in a middle-income country, demonstrates that institutional obstacles to improving hand hygiene can be substantial, and that without additional resources attempts to change behavior using the WHO’s multimodal strategy are not guaranteed to succeed. While we do not have a good idea of how widespread such refractoriness is, the strong evidence of publication bias in the literature and the high frequency of reported compliance below 20% suggest that our findings may be representative, at least for a substantial subset of hospitals in LMICs.

We believe the vital role that hand hygiene plays in preventing infections and controlling antimicrobial resistance should make it a public health priority to establish country-level hand hygiene programs that ensure acceptable hand hygiene compliance in every hospital. This is likely to require independent hand hygiene audits and appropriately-resourced nationally coordinated hand hygiene campaigns based on the WHO strategy or evidence-based extensions of this. Such national campaigns have had remarkable success in high income countries, and recent analysis has shown that unless baseline compliance is already very high, such investment in hand hygiene will almost certainly be cost-effective (and often cost-saving) in lower income settings. Country-wide roll-out out of such campaigns to hospitals in a stepped wedge manner would also enable the generation of robust evidence concerning the effectiveness and cost-effectiveness of the intervention.

## Supporting information

CONSORT checklist for stepped wedge RCT

ICMJE DISCLOSURE FORM

## Data Availability

All data produced in the present study are available upon reasonable request to the authors

## Contributors

BSC and DL conceptualized the project and designed the study. BSC supervised the project, acquired funding, and is the manuscript’s guarantor. MH, PS, SN, collected and curated the data. MH cleaned the data. MH and CL performed the formal data analysis. MH and CL produced the figures. MH and CL drafted the manuscript. All authors interpreted the data and critically revised the manuscript for intellectual content. All authors had full access to all the data in the study. All authors, read and approved the final manuscript.

